# “Enhancing Nursing Education through Mobile Health Clinics: Aligning AACN Core Competencies with Pediatric Clinical Experiences in Rural and Indigenous Communities”

**DOI:** 10.1101/2024.08.21.24312378

**Authors:** Shelly Hogan, Madeline Metcalf, Ann Galloway, Nicole Krueger, Laura Larsson

**Affiliations:** Mark & Robyn Jones College of Nursing, Montana State University, Bozeman, MT, USA

**Keywords:** Nursing, Nurse Education, Indigenous Health, Rural Health, Culturally Responsive Care, Public Health Nursing, Nurse Competency

## Abstract

**Aim:** This study provides insights into student perceptions of a hands-on, interprofessional pediatric clinical experience within Montana’s tribal communities through the Mobile Health Training Program (MHTP). Specifically, it explores how the MHTP aligns with AACN core competencies and evaluates the impact of these practical experiences on nursing students’ competencies and readiness.

**Background:** A well-prepared nursing workforce is crucial for addressing the healthcare needs of rural, underserved, and Indigenous communities in the United States. Montana’s Indigenous communities face significant systemic economic and social challenges that limit access to healthcare services and contribute to a disproportionate disease burden. Over the past two years, the MHTP at a land grant university in the western U.S. conducted preschool health screening clinics for children aged 0-5 at four partner Tribal nations in Montana.

**Design:** This qualitative study captures and analyzes the experiences and perceptions of undergraduate nursing students participating in the MHTP at a land grant university in the western U.S. Data were collected through post-MHTP clinic surveys administered through Qualtrics, after the completion of each 2-3 day MHTP clinic.

**Methods:** A total of 160 nursing undergraduate nursing students completed post-clinical surveys between September 2022 and May 2024. Using inductive and deductive coding approaches, survey responses were analyzed qualitatively to identify themes related to rural healthcare, culturally responsive care, exposure to diverse communities, and professional growth.

**Results:** The MHTP experience was found to be valuable by 95% of students. Four main themes emerged: insight into rural healthcare, practicing culturally responsive care, exposure to diverse communities, and professional growth. American Indian/Alaska Native student perspectives highlighted the need for culturally reflective opportunities for all students. The MHTP effectively aligned with AACN competencies, enhancing skills in patient-centered care, cultural competence and humility, interprofessional collaboration, and systems-based practice.

**Conclusion:** The MHTP represents a contemporary and practical model in nursing education by integrating mobile, immersive, and patient-focused clinical experiences, while also providing students an opportunity to engage with high priority communities. Students valued working with rural and Indigenous populations, recognizing the critical need for culturally responsive care and professional development. This study contributes to the limited literature on rural and Tribal clinical placements, highlighting the importance of balancing simulation with direct clinical practice. The MHTP effectively equips nursing students to navigate the complexities of healthcare systems in rural and underserved settings and advancing nursing education to address workforce needs in these areas.

## INTRODUCTION

The American Association of Colleges of Nursing (AACN) outlined core competencies through *“The Essentials: Core Competencies for Professional Nursing Education,”* a framework designed to prepare a competent nursing workforce.(American Association of the Colleges of Nursing, 2021) This framework is integral to ensuring that nursing education meets the practical demands and healthcare needs of diverse populations, particularly in rural and underserved communities. Embedded in AACN *Essentials* is competency-based education (CBE), a pedagogical approach that focuses on mastering key competencies rather than traditional input-based measures, emphasizing outcomes and practical application.(American Association of the Colleges of Nursing, 2021; National Academies of Sciences, Engineering, and Medicine; National Academy of Medicine; Committee on the Future of Nursing 2020–2030, 2021) Similarly, the American Public Health Association (APHA) emphasizes the importance of a well-prepared and diverse health workforce in strengthening public health systems. The APHA defines ten Essential Public Health Services that impact the strength and capacity of a public health system, including the preparation and diversity of the health workforce.(*10 Essential Public Health Services*, n.d.) Despite the importance of these frameworks, nursing education has often been centered in urban areas, leaving a gap in preparation for rural and Indigenous healthcare settings.

In healthcare education, CBE is reflected in curricula, coursework, and clinical practices designed to foster learning and ongoing professional development. Students are expected to engage in continuous performance assessment and reflection to enhance their skills. The AACN Core CBE Domains, outlined in *“The Essentials: Core Competencies for Professional Nursing Education,”* provide a comprehensive framework for advancing nursing practice.(American Association of the Colleges of Nursing, 2021) These domains include: 1) Knowledge for Nursing Practice, focusing on the integration and application of disciplinary knowledge; 2) Person-Centered Care, emphasizing holistic and evidence-based approaches; 3) Population Health, which involves collaborative efforts to achieve equitable health outcomes; 4) Scholarship, the generation and dissemination of nursing knowledge; 5) Quality and Safety, applying improvement science to enhance safety and quality; 6) Interprofessional Partnerships, underscoring collaboration with other professionals; 7) Systems-Based Practice, navigating complex healthcare systems; 8) Informatics and Technology, employing data-driven methods; 9) Professionalism, developing a professional identity based on accountability; and 10) Personal, Professional, and Leadership Development, encouraging lifelong learning and resilience.(American Association of the Colleges of Nursing, 2021) Together, these domains ensure that nursing education addresses current healthcare demands and prepares practitioners for future challenges, especially in high priority populations.

Indigenous, isolated, and rural communities often face unique barriers to healthcare, including geographic isolation, limited transportation, economic insecurity, lack of health education resources, higher rates of uninsurance, lack of primary care and specialty providers, insensitive cross-cultural communication, and health system trust concerns.(Cromer et al., 2019) Montana, a rural state in the U.S. Mountain West, is home to 12 tribal nations residing on seven federally recognized reservations. Montana’s Indigenous people make up about 6.5% of the state’s population, representing the second-highest racial group.(*Montana - 2021 - III.B. Overview of the State*, n.d.) Tribal communities shape the vitality and culture of Montana; however, Indigenous populations have historically been medically underserved and experience disproportionate challenges in accessing health services. Specifically, 50 out of 56 counties in Montana are categorized as primary care provider shortage areas and 44 out of 56 are dental provider shortage areas designated by the U.S. Health Resources and Services Administration (HRSA).(*Montana Health Professional Shortage Area (HPSA) Designations*, n.d.; *Rural Healthcare Workforce Overview - Rural Health Information Hub*, n.d.) Nationally, research underscores the need to increase health workforce capacity in these communities, and Montana is no exception. Expanding the health workforce in rural and underserved areas is crucial, and nursing educational programs will play a key role in preparing students for careers in these communities.

In response to these challenges, the Mobile Health Training Program (MHTP) at Montana State University (MSU) offers a model for addressing healthcare needs in rural and isolated communities. Since its inception in 2018, the MHTP has assembled interprofessional teams to provide essential healthcare services to four Tribal partner nations. This program features undergraduate student-led mobile clinics staffed by an interprofessional team, including nursing faculty, nursing student clinicians, dental hygienists, dentists, optometrist, audiologist, and a developmental therapist. The MHTP delivers a range of health and educational services to high priority communities, such as screenings, preventive treatments, specialty referrals, nutrition and oral health education, and case management. (Larsson & Hodgson, 2023) Through their involvement in the MHTP, nursing students gain valuable hands-on pediatric clinical experience, develop interprofessional collaboration skills, and build a solid foundation in delivering culturally responsive care. Each year, the MHTP conducts at least 10 clinics in partnership with Montana’s tribal communities, with staff traveling an average of 204 miles to each site. Since 2019, 658 undergraduate nursing students have participated in the program, completing clinical hours at partner nations such as the Amskapi Pikuni (Blackfeet), Selis, Ksanka, and Qlispe (Flathead), Tsististas and Suhtaio (Northern Cheyenne), and Apsaalooke (Crow) tribes.

While there is a growing body of nursing education literature focusing on undergraduate student exposure to service learning, social determinants of health, cross-cultural experiences, and rural healthcare simulation exercises, (Alexander-Ruff & Kinion, 2018; Eddie et al., 2023; Hendrickx et al., 2014; Liu et al., 2024; Mattingly, 2021; Thornton & Persaud, 2018) our research addresses a distinct gap. This study provides insights into student perceptions of a hands-on, interprofessional pediatric clinical experience within Montana’s tribal communities through the Mobile Health Training Program (MHTP). Specifically, it explores how the MHTP aligns with AACN core competencies and evaluates the impact of these practical experiences on nursing students’ competencies and readiness. By examining how the MHTP supports and reflects AACN’s essential competencies, this article aims to demonstrate the value of immersive clinical training in preparing nursing students to meet the diverse and complex needs of rural and indigenous healthcare settings.

## METHODS

### Study Design

This qualitative study aimed to capture and analyze the experiences and perceptions of undergraduate nursing students participating in the MHTP at a land grant university in the western U.S. Data were collected through post-MHTP clinic surveys administered through Qualtrics, after the completion of each 2-3 day MHTP clinic. The survey period spanned from September 2022 through May 2024, including both fall and spring clinic sessions. During this timeframe, the MHTP provided comprehensive health services to preschool children aged 0-5 years within the Blackfeet, Flathead, Northern Cheyenne, and Crow tribes. The student-led mobile clinics provide interprofessional health services including dental, vision, hearing, developmental screenings, as well as preventive care and case management for a total of 2,036 Indigenous children. In addition to essential health screen services (i.e. height, weight, blood pressure, vision, hearing, blood lead, blood hemoglobin, and developmental), a notable component of the program involved a focus on oral health. Under the supervision of dental hygienists nursing students assisted in applying fluoride varnish applications, observing Silver Diamine Fluoride (SDF) treatments and dental sealant applications. Through these clinics, nursing students gained valuable exposure to specialized care, case management, and various aspects of service delivery, contributing to improved health outcomes for the pre-school communities served.

### Setting

Montana’s Indigenous communities face significant systemic economic and social challenges that limit access to healthcare services and contribute to a disproportionate disease burden.(*Montana - 2021 - III.B. Overview of the State*, n.d.) Nearly 60% of Indigenous people in Montana reside on tribal lands, with all Montana reservations classified as mental health, primary care, and dental provider shortage areas.(*Montana Health Professional Shortage Area (HPSA) Designations*, n.d.) Compared to the U.S. national median age of 32.8 years, the Indigenous population in Montana is notably younger, with a median age of 27.3 years, emphasizing the need for targeted healthcare interventions for younger populations.(Holom, 2020) The Mark and Robyn Jones College of Nursing (MRJCON) at Montana State University, the state’s Land Grant institution, is dedicated to training nurses, nurse educators, nurse midwives, and nurse practitioners to serve Montana’s urban, rural, frontier, and tribal communities. With an annual undergraduate nursing enrollment of 328, the MRJCON operates five nursing campuses (Bozeman, Billings, Missoula, Great Falls, and Kalispell), providing students with diverse clinical experiences across the state.

### Participants

Participants included traditional and accelerated Bachelor of Science in Nursing (BSN) students from MSU who completed clinical rotations with the MHTP during the two-year study period (September 2022 – May 2024). A total of 160 survey responses were collected, representing 73.7% of the students involved in the program over the past two years (160/217). Of these respondents, 17 identified as American Indian/Alaska Native (AI/AN), making up 10.6% of the surveyed group.

### Data Collection Procedures

Following their MHTP clinical experience, students were invited via email to complete a survey administered through Qualtrics Software (Silver Lake, Seattle, WA, USA).(*Who Is Qualtrics?*, n.d.) Professional nursing faculty informed the students about the survey and that their responses could be published and used to improve the MHTP. An electronic informed consent form was completed before starting the survey. The survey, developed by the research team, included a combination of multiple-choice, short-answer, and satisfaction questions addressing various aspects of the clinical experience. Short answer prompts allowed students to offer their perspective on the value of the practical experience, suggest improvement, provide examples of clinical engagement, share surprising or unexpected events, and highlight particularly affirming encounters. Students received a gift card upon completing the survey.

### Analysis

Upon completion of the 2022-2024 Mobile Health Training Program (MHTP) at the four pediatric clinic sites, 160 complete survey responses were collected via Qualtrics. The qualitative data were analyzed using both inductive and deductive coding approaches to identify recurring themes and prevalent insights. The preliminary qualitative analysis was conducted by the primary analyst (SH), who developed a codebook based on survey categories and emerging themes to guide the analysis and results presentation. A secondary analyst (MM) reviewed the data to extract additional insights and ensure consistency between student feedback and identified themes. To compare perceptions between self-identifying AI/AN and non-AI/AN students, data were further filtered between these groups. No major discrepancies were found between the two analysts in terms of theme definitions or quote inclusion. Finally, based on discussion between the research team, the themes presented within this article were aligned with AACN core CBE Domains.

### Ethical Considerations

The study’s survey data collection and storage procedures received approval from the university’s Institutional Review Board (IRB) before initiation (IRB 2019-759-LL061419).

## RESULTS

A total of 160 post-clinical experience surveys were completed by undergraduate nursing students participating in the MHTP between September 2022 and May 2024. An overwhelming 95% (n=152) of students found the MHTP experience valuable, highlighting the importance of providing high quality clinical experiences where students can synthesize principles from pediatrics and community health, practice providing culturally responsive care with diverse populations, and gain first-hand experience with challenges faced by rural and indigenous populations. The following qualitative results detail student perceptions within four identified themes and note insights from AI/AN nursing students.

### Insight into Rural Healthcare

Students reported that their MHTP clinical experience provided a deeper understanding of healthcare in rural communities. They described gaining a *“first-hand”* perspective of rural health issues, which included financial constraints, geographic isolation, and general lack of access to healthcare providers. Many nursing students recognized the equity-building of providing healthcare in these communities. One student emphasized the importance of equitable care, stating, *“Interacting with people who are not in the hospital is just as important for those who are; everyone deserves quality health care!”*

### Practicing Culturally Responsive Care

Nursing students reported that their MHTP experience with tribal partners helped them “*understand the need to tailor care*” and “*develop cultural humility*.” They viewed cultural humility as an ongoing journey rather than a measurable endpoint or achievement. For instance, one student mentioned, “*It was also my first time on a reservation and being a future nurse in Montana, understanding their culture better is crucial!*” Students emphasized the value of cross-cultural clinical opportunities, with one stating, “*Having to practice cultural competence is more valuable than I can put into words*.”

### Exposure to Diverse Communities

Students valued the opportunity to work with communities often not included in traditional clinical placements. They noted that seeing different communities highlighted how environmental factors and resource access affect health. One student noted, “*You get to see communities you wouldn’t typically see and how the environment and access to resources affect a community! Rural health isn’t included in our clinical schedule, and I think it should be so people can see the differences.*” Another student shared that the experience allowed them to “*experience diversity in a different light*.” While students valued the experience, some noted the need for systemic change to address broader issues in rural and indigenous healthcare, with one stating, “*I think learning how to conduct healthcare on a population that we don’t see very much is very helpful… However, I do feel a bit like it is a Band-Aid over a bigger problem, but I think the role we play is important and helpful*.”

### Professional Growth

Nursing students noted how their MHTP experience contributed to their professional growth and readiness for a career in nursing. Students reported increased confidence and enhanced skills. One student shared, *“It helped instill confidence and the courage to take charge in assessing the children and I feel gave me more confidence in future interactions with pediatric patients.”* Another noted, *“By going out to this clinical, I have strengthened my communication skills as well as my nursing skills and will take them forward with me in my career.”* Overall, student responses suggest that they valued their *“hands-on”* MHTP experience as an opportunity for professional growth.

### AI/AN Student Perspectives

One of the main themes voiced by American Indian/Alaska Native (AI/AN) students was their surprise at how little their peers knew about healthcare and culture in tribal communities. Despite this, they were optimistic that the experience would enhance their peers’ ability to deliver quality healthcare. One AI/AN student remarked, *“I feel like what surprised me the most is that some of my classmates have never seen the rez [reservation] or even been on the rez [reservation]… I think it was a good experience for them to become a little more well-rounded for a lack of better words. The more we can all experience as healthcare workers - the better the care we can give. It’s good to see where people come from and I’m glad they got the chance to see that.”* Other AI/AN students noted, “*serving my community*” and *“seeing how supportive my community is”* as affirming experiences, underscoring the importance of culturally reflective clinical opportunities for all students.

### Alignment with AACN Essential Core Competencies

In the AACN “*Essentials: Core Competencies for Professional Nursing Education*,” the ten domains provide a comprehensive framework for developing nursing practice. The results of this study align with several domains of the AACN Essential Core Competencies for Professional Nursing Education. This alignment demonstrates that the MHTP serves as a competency-based education (CBE) model that prepares nursing students to work with diverse and high-priority populations (Table 1).

**Table 1:**
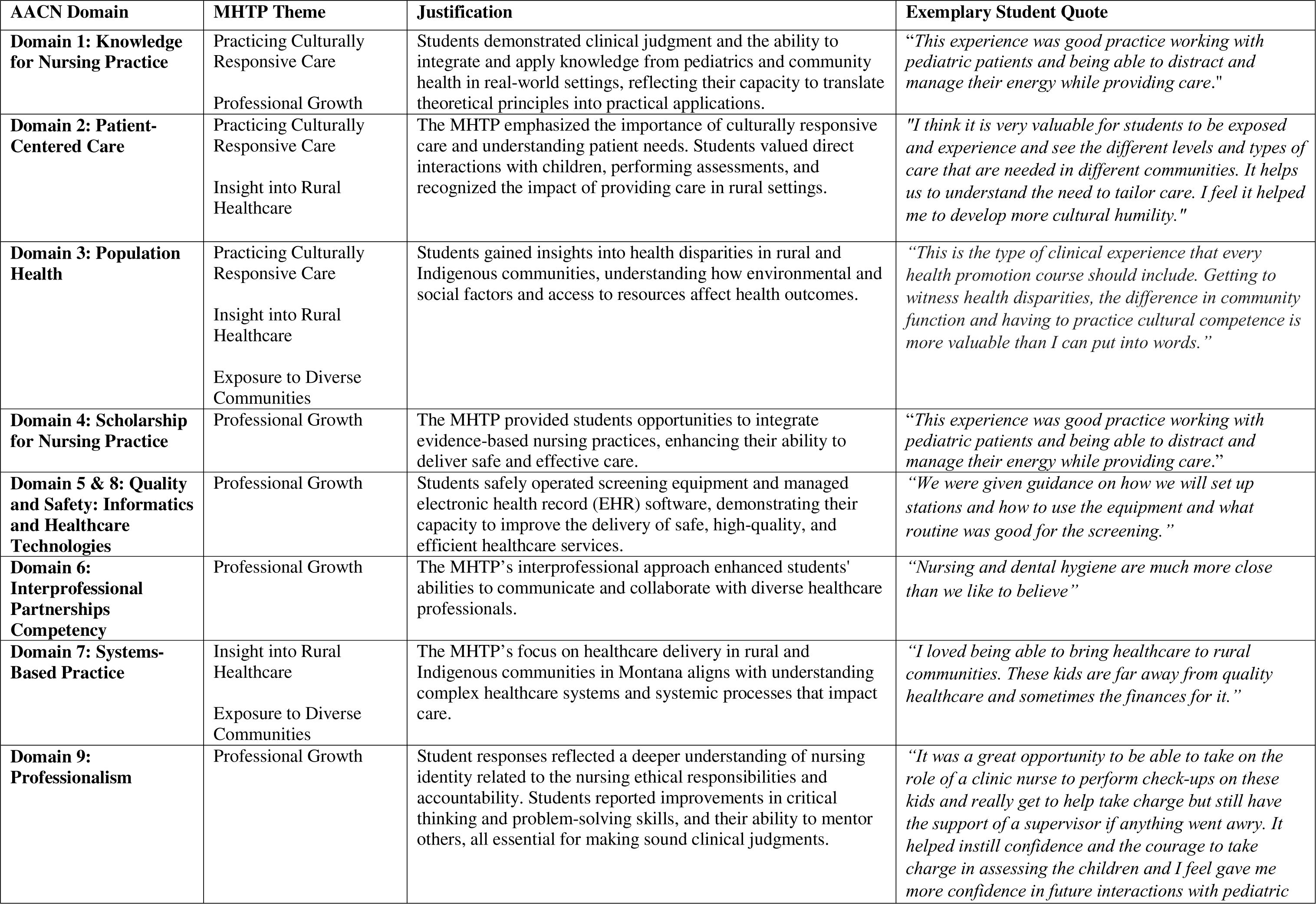
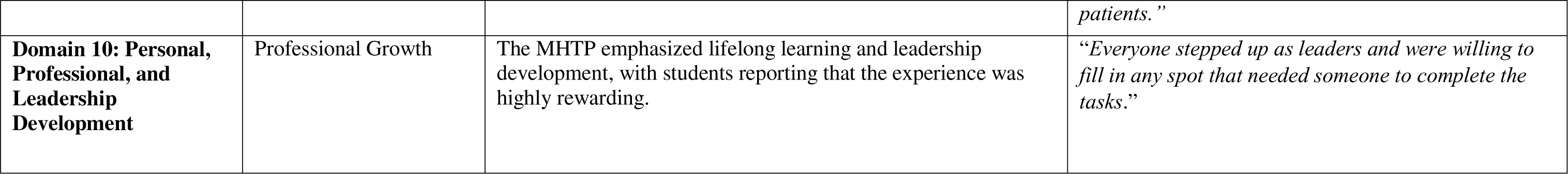
Select Quotes by AACN Competency Domains and Themes.

By integrating the ten core AACN competencies, the MHTP provides an effective model for preparing nursing students to meet the evolving demands of practice, especially in rural, underserved, and diverse populations. This program incorporates CBE domains such as cultural sensitivity, interprofessional collaboration, and professional development. It equips students to deliver high-quality and competent care across a range of contemporary challenging settings, addressing the complex needs of diverse communities.

## DISCUSSION

This study provides beneficial insights into nursing student perceptions of an in-person mobile clinical experience with tribal nations in Montana. To our knowledge, this is the first study to align the AACN core competencies for nursing education with student perceptions of a rural, indigenous clinical experience, contributing to the limited literature on the value of these clinical placements for undergraduate nursing students. The findings highlight several key themes: the need for culturally responsive care, the advantages of firsthand exposure to rural health and diverse communities, and the development of professional nursing competencies.

One of the most frequently mentioned benefits of the MHTP was the enhancement of students’ culturally responsive care skills. Students recognized that providing culturally safe care involves ongoing self-awareness and humility. This aligns with the growing emphasis in medical, public health, and nursing curricula on preparing the workforce to deliver culturally competent care.(*Establishing a Culturally Competent Master’s and Doctorally Prepared Nursing Workforce - ANA Position Statement*, 2018; *Support for Culturally and Linguistically Appropriate Services in Health and Mental Health Care*, n.d.; *The AMA’s 2024—2025 Strategic Plan to Advance Health Equity*, 2024) In states like Montana, where nearly 85% of the population identify as white, it is challenging to find clinical opportunities that allow students to apply cultural safety principles effectively.(Bureau, n.d.) Indigenous students often face the expectation of providing cross-cultural care in a variety of settings. Our findings emphasize the need for nursing programs to create diverse clinical placements that not only provide practical experience but also a sense of fit and belonging for all students. As such, we advocate for programs that offer both safe spaces for practicing cross-cultural care and empower students to provide competent and culturally inclusive care.

Students highly valued the opportunity to engage with communities that are not typically included in their traditional urban clinical schedules. Research highlights the importance of addressing nursing workforce shortages in rural areas and the benefits of early exposure to rural nursing opportunities.(Terry et al., 2024) Studies have shown that rural placements significantly influence nursing students’ career considerations, especially for those who complete multiple rural clinical rotations.(Campbell et al., 2021) Evidence from these studies suggests that exposure to rural nursing opportunities should occur early and often in a student’s course of study. To increase student exposure, nursing curricula has implemented simulation exercises centered around rural healthcare delivery and sociocultural considerations when working in rural, underserved, or Indigenous communities.(Eddie et al., 2023; Hendrickx et al., 2014) Simulation exercises are useful in offering practice with nursing skills while simultaneously limiting the real logistical challenges of travel, minimal placements, and outreach in these communities. However, our data underscore the students’ preference for hands-on clinical experiences. Successful examples of these experiences are detailed in research conducted by Alexander-Ruff and Mattingly, which illustrate indigenous clinical and service-learning opportunities that facilitate culturally safe learning with nursing students in Montana and South Dakota.(Alexander-Ruff & Kinion, 2018; Mattingly, 2021) Therefore, we recommend that future research and educational curriculum strategies integrate both simulation and real-world clinical opportunities to enhance students’ skills and understanding of rural health nursing, culturally responsive care, and social determinants of health.

The MHTP presents an applicable model for overcoming barriers to rural placements. Its unique approach includes student-led mobile clinics made up of an interprofessional team that bring healthcare directly to underserved communities, reducing logistical challenges and increasing accessibility. The program’s design highlights its potential as a scalable solution for other nursing education programs facing similar challenges. Specific strategies employed by the MHTP include: 1) Student-Led Mobile Clinics - These clinics effectively bridge the gap between healthcare providers and remote communities, making it easier for students to engage with underserved populations while being educated and exposed to the essential AACN competences domains. 2) Communication and Relationship Building - Establishing strong partnerships with indigenous communities before the clinical experience begins ensures that the program is personalized to the specific healthcare needs of these communities and facilitates more immersive and impactful experiences for students. and 3) Community-Focused Experiences - By prioritizing community involvement and responsiveness, the MHTP fosters deeper connections between students and the communities they serve, enhancing the educational value and relevance of the clinical placements. These strategies not only address logistical and accessibility issues but also demonstrate how such models can be customized and scaled to meet the needs of both students and high priority communities and at the same time align with the AACN domain competencies. The success of the MHTP in providing meaningful, community-centered experiences highlights its potential as a blueprint for other nursing education programs aiming to improve rural clinical placements.

Although the data we present offers insight into the value, successes, and opportunities of a rural, indigenous, and pediatric clinical opportunity for undergraduate nursing students, our study contains limitations. As a qualitative study, the findings are not intended to be generalizable but provide insight into the experiences of nursing students who participated in the MHTP from September 2022 to May 2024. The study does not capture the perspectives of students involved in the program before this period, which could have offered different insights. Additionally, our post-survey prompts students to report their knowledge, ability, and comfort both before and after the clinical experience on a variety of factors, and thus the self-reported data may be subject to recall bias. While we did not formally assess inter-rater reliability, routine discussions between the qualitative evaluation team ensured consistency in theme identification. Despite these limitations, the findings provide valuable insights into the benefits and challenges of rural and indigenous clinical placements.

## CONCLUSION

This article illustrates how the MHTP integrates AACN competencies, offering a practical example of how educational programs can adapt to nursing pedagogical frameworks and enhance student learning outcomes. We highlight nursing student perspectives of a clinical opportunity in rural and indigenous communities. We recommend that 1) supplementary clinical programs thoughtfully incorporate diverse experiences that empower professional development for both minority and majority culture students and 2) research and education programs explore a mixture of simulation and hands-on clinical opportunities for increasing exposure and awareness of rural nursing careers. Additionally, the broader implications related to nursing education is better understanding and showing how similar models could be applied or adapted in other settings or programs. Potential areas for further research based on the study findings could include exploring the long-term impact of the MHTP on career outcomes for nursing students or assessing the effectiveness of similar programs in other regions and populations. In conclusion, the MHTP exemplifies how contemporary, competency-based, hands-on training can effectively prepare nursing students for the unique challenges of rural and indigenous healthcare settings. By aligning with AACN competencies and addressing critical healthcare needs, MHTP help advance nursing education and address workforce disparities in underserved communities.

## Data Availability

All data produced in the present study are available upon reasonable request to the authors

## Acknowledgements

Authors wish to acknowledge our tribal partners, the Amskapi Pikuni, Apsaalooke, Selis, Kanka, Qlispe, Aaniiih, Assiniboine, Tsististas and Suhtaio at four sites who invite us to bring students, host the clinics, and facilitate interprofessional and intercultural clinical experiences.

## Author Contributions

Conceptualization: SH, MM

Methodology: LL, SH, MM

Formal Analysis and Investigation: SH, MM

Data Collection: SH, LL, NK

Writing – Original Draft Preparation: SH, MM

Writing – Review and Editing: MM, SH, LL, NK, AG

Funding Acquisition: LL

Supervision: LL, SH

